# Correlates of suicidality in men with psychogenic sexual dysfunction

**DOI:** 10.1101/2020.06.03.20121822

**Authors:** Ravi Philip Rajkumar

## Abstract

**Background:** Psychogenic sexual disorders in men, such as erectile dysfunction and premature ejaculation, are often associated with anxiety and depression. However, no study has systematically examined the link between these disorders and suicidality.

**Aim:** To assess the frequency and correlates of suicidal ideas and behaviour in men with a primary diagnosis of psychogenic sexual dysfunction.

**Method:** The current study is a retrospective chart review of 64 Indian men with psychogenic sexual dysfunction, presenting to our clinic in the period 2013–14. Demographic and clinical variables associated with the presence of suicidal ideation or attempts in these patients were examined.

**Results:** Nine (14.1%) patients experienced suicidal ideation related to their disorder, and four (6.3%) had made a suicide attempt. Depression, childhood adversity, and a family history of alcoholism were associated with suicidal ideation, while ongoing stressful life events were associated with suicide attempts.

**Conclusions:** Assessment of suicidality should be part of the routine evaluation of men with sexual dysfunction, particularly when other risk factors are present.

## Introduction

Erectile dysfunction (ED) and premature ejaculation (PE) are the commonest forms of male sexual dysfunction (1, 2). When these disorders occur in the absence of a clear medical cause, or in the presence of clear psychosocial factors related to their onset, they are said to be psychogenic (3). These psychosocial factors include childhood adversity, relationship conflicts, stressful life events and anxiety related to sexual performance (4, 5).

Comorbid depressive and anxiety disorders are common in these patients, but frequently go undiagnosed (6, 7). Patients with ED (8) and PE (9) may also have symptoms of anxiety and depression not fulfilling the criteria for a syndromal diagnosis. The relationship between depression and sexual dysfunction is complex, as either condition can increase the risk of developing the other (10, 11).

As these disorders are associated with an increased risk of suicide (12), we would expect to find elevated rates of suicidal ideation and behaviour in men with sexual dysfunction and comorbid depression or anxiety. This may be more likely in traditional societies where an inability to “perform” sexually is viewed as a failure of masculinity (13). However, the association between suicidality and sexual dysfunction has not been systematically explored. The current study is a preliminary attempt at addressing this issue. We carried out a retrospective assessment of the frequency and correlates of suicidal ideas and attempts in men primarily seeking help for psychogenic sexual dysfunction.

## Methodology

### Patient population

The case records of 64 men with a primary diagnosis of psychogenic erectile dysfunction or premature ejaculation, seeking help at a clinic for psychosexual disorders in a general hospital in South India in the period 2012–2014, were reviewed. Patients are generally referred to the clinic by specialty departments, such as urology or general surgery, after an evaluation for organic causes of sexual dysfunction.

### Assessments

All patients attending the clinic are interviewed by a psychiatry resident using a semi-structured interview schedule which records details of sexual dysfunction, comorbid psychiatric disorders, family histories of psychiatric and substance use disorder, and associated psychosocial variables such as childhood adversity, marital disharmony and other ongoing stressors. Information on suicidal ideation or attempts related to sexual dysfunction is also recorded. All psychiatric diagnoses are confirmed by the consultant in charge of the clinic (the author) and are made using the World Health Organization’s International Classification of Diseases, Tenth Edition (ICD-10) clinical descriptions and diagnostic guidelines. This study was conducted in accordance with the guidelines of the institute’s Review Board, which permits chart reviews of clinical data as long as confidentiality and anonymity are not violated.

### Data analysis

Patients with and without suicidal ideation and behaviour were compared in terms of their demographic and clinical profile, and binary logistic regression analyses were carried out to identify variables associated with these outcomes. The chi-square test or Fisher’s exact test was used to compare categorical variables, and the independent samples *t* test or the Mann-Whitney *U* test were used to compare continuous data. All univariate analyses were two-tailed except where specified in the text, and a value of *p* < 0.05 was considered significant.

## Results

### Description of the sample

We obtained information on 64 men with a presenting diagnosis of ED or PE. The mean age of these patients was 31.3 ± 6.2 years (range 22 to 46 years). 30 men were single, 31 married, one divorced and two widowed. Thirty-seven of these men (57.8%) had a diagnosis of premature ejaculation, and forty-four (68.8%) had erectile dysfunction; 17 patients (26.6%) had both PE and ED. None of these patients had been diagnosed with, or were being treated for, a psychiatric disorder at the time of their assessment.

### Frequency of suicidal ideation and behaviour

Suicidal ideation related to current sexual problems was documented in nine men (14.3%), and four of these men (44.4%) had made a suicide attempt. The methods used by these men included hanging in two cases, wrist-slashing in one case, and a medication overdose (paracetamol) in one case. The latter two patients were treated in local hospitals for their attempts, but none of the four specifically sought psychiatric help for suicidal ideation. None of these patients reported intentions of making a second attempt.

### Correlates of suicidal ideation

When comparing men with (SI+) and without (SI-) suicidal ideation **(Table 1)**, we found no significant difference in terms of the type of sexual dysfunction. PE was slightly less common in the SI+ men, but this was not statistically significant.

The presence of a comorbid depressive disorder, particularly major depression, was significantly higher in the SI+ group (*p* = 0.01, Fisher’s exact test). In contrast, neither anxiety disorders nor sexual performance anxiety were associated with suicidal ideation. Parental marital discord during patients’ childhood (*p* = 0.04, Fisher’s exact test) and a family history of alcoholism in a first-degree relative (*p* = 0.016, Fisher’s exact test) were significantly more frequent in the SI+ group. Overall, adverse events during childhood were more common in the SI+ group (Mann-Whitney *U* = 363.5; *p* = 0.015).

### Correlates of suicide attempts

When men who had made a suicide attempt (*n* = 4) were compared to those with no current suicidality (*n* = 55), only two variables differed significantly: comorbid depressive disorders were more common (*p* = 0.048, Fisher’s exact test) and the presence of an ongoing stressor (*p* = 0.024, Fisher’s exact test). Stressors reported by these men included: having a child with intellectual disability (one case), the recent break-up of a romantic relationship (one case), and conflicts with the patient’s own father (one case).

## Discussion

Suicidal ideation was seen in 14% of patients who sought help for sexual dysfunction. However, only a minority of those with such ideation made a suicide attempt. This is consistent with evidence suggesting that additional factors, such as personality and sociodemographic variables, mediate the transition from suicidal ideas to actual attempts (14, 15).

The type of sexual dysfunction experienced by the men in our study did not show any association with suicidality. This is consistent with evidence that both erectile dysfunction (16) and premature ejaculation (17) are associated with low self-esteem, which is associated with an increased risk of suicide (18). Our findings suggest that the presence rather than the exact nature of sexual dysfunction causes emotional distress and triggers suicidal ideation.

Comorbid depression, particularly major depression, was a predictor of suicidality. Suicidal ideation was more strongly associated with major depression than with dysthymia. This is consistent with the recent finding that suicidal ideation is prominent in more severe forms of depression (19).

Childhood adversity, particularly in the form of parental marital disharmony and conflict, was more common in patients with suicidal ideation. This association was not mediated by comorbid depression. Our results are in line with earlier findings on the relationship between childhood adversity and suicidality in later life (20, 21), including an association between parental separation and suicidality (22).

A family history of alcoholism in a first-degree relative was associated with suicidal ideation, and remained significant even in the logistic regression analysis. Such an association may reflect a possible shared genetic vulnerability for alcoholism and suicidality (23, 24). However, it may also be related to the early life adversities experienced by patients growing up with an alcoholic family member (25).

The presence of stressful life events was a stronger predictor of suicide attempts than of suicidal ideation. This is consistent with models of suicidality as a process or continuum (26) in which distal risk factors, such as genetic vulnerability or early life adversity, interact with proximal risk factors such as current mental illness or stressors. Applying such a model to our sample, distal risk factors such as a family history of alcoholism and adverse childhood experiences may have created a general vulnerability to suicide. Entry into the suicidal process may have been triggered by sexual dysfunction, causing low self-esteem (18) and leading to suicidal ideation, which would further lead to suicidal behavior in those experiencing stressful life events.

Our study is subject to certain important limitations. First, due to the retrospective, chart-based design, it is subject to problems with missing information. Second, suicidal ideas and attempts were assessed as simply present or absent; no structured instrument was used to assess suicide intent or other psychological variables of interest. Third, due to the small absolute number of patients with suicidal ideation or behaviour, the study may have been underpowered. Fourth, our patients were recruited from a tertiary care hospital, and may not accurately represent men with sexual dysfunction in the general population. Fifth, all diagnoses were made using a clinical interview alone; subsyndromal depression and anxiety were not assessed using a rating scale. Finally, information on childhood adversity was obtained from patients using simple yes/no questions, and could be corroborated by family members in only some cases; thus, this information may be subject to recall and response bias.

Despite these limitations, our study is the first of its kind to examine the frequency of suicidal ideas and behavior in men with sexual dysfunction. From a clinical perspective, our study highlights the need to screen for and manage suicidal ideas and behaviour in men with a primary complaint of sexual dysfunction, particularly when other risk factors – such as depression, ongoing negative life events, or early life adversity – are also present.

## Clinical Points

1. A substantial number of men with sexual dysfunction experience suicidal ideas.
2. Depression and childhood adversity may increase the risk of suicidal ideation in these patients.
3. Men with sexual dysfunction should be carefully assessed for co-occurring depression, anxiety and suicidality.

## Data Availability

Data for this study will be available on request.

